# Estimating thresholds for risk of cannabis use disorder using standard THC units

**DOI:** 10.1101/2025.05.21.25328059

**Authors:** Rachel Lees Thorne, Will Lawn, Kat Petrilli, Katie Trinci, Anya Borissova, Shelan Ofori, Claire Mokrysz, H Valerie Curran, Lindsey A Hines, Tom P Freeman

## Abstract

**Background & Aims:** Lower risk guidelines for safer levels of cannabis use could help to reduce the health burden posed by cannabis use disorder (CUD). We aimed to estimate risk thresholds for CUD based on delta-9-tetrahydrocannabinol (THC) consumption using standard THC units (1 unit = 5mg THC).

**Design:** Data from the CannTeen study, a longitudinal observational study consisting of five assessments over a 12-month period.

**Setting:** London, UK.

**Participants:** Participants were *n* = 65 adults aged 26-29 (46% female, 70.77% white ethnicity) and *n* = 85 adolescents aged 16-17 (56% female, 65.48% white ethnicity). All participants reported at least one use of cannabis during the 12-month study period and completed an assessment of DSM-5 CUD symptoms at the final follow up.

**Measurements:** Mean weekly standard THC units were estimated using the Enhanced Cannabis Timeline Followback, a comprehensive assessment of quantity, frequency and potency of cannabis consumed. This was administered at 3-month intervals and averaged over a 12-month period. Past 12-month diagnosis of DSM-5 CUD was assessed at the final follow-up. Receiver operating characteristic curve models estimated the extent to which weekly standard THC unit consumption could discriminate no CUD from any CUD (mild, moderate, or severe), and no CUD from moderate/severe CUD, in adults and adolescents separately. Risk thresholds were selected based on cut-offs that maximised sensitivity and specificity.

**Findings:** Discrimination accuracy of weekly standard THC units on CUD was good, with area under the curve > 0.70 for all models. Optimal cut-offs for risk of any CUD (versus no CUD) were 8.26 units per week for adults and 6.04 units per week for adolescents. For risk of moderate/severe CUD (versus no CUD) optimal cut-offs were 13.44 units per week for adults and 6.45 units per week for adolescents.

**Conclusions:** Standard THC units show good discrimination accuracy of CUD at different severities and in different age groups. Safer levels of cannabis use, defined by low weekly standard THC unit consumption, could be recommended in lower risk cannabis use guidelines.

## INTRODUCTION

An estimated 22% of people who use cannabis will develop cannabis use disorder (CUD), (1). CUD refers to a pattern of cannabis use that causes clinically significant impairment or distress. Symptoms include using cannabis for longer than intended, tolerance to the effects of cannabis, not meeting obligations at work/school due to cannabis use, and spending a lot of time getting, using or recovering from the effects of cannabis (2). Cannabis use is responsible for 35% of EU drug treatment entrants and is cited as a problem drug by 87% of under-18’s in drug treatment in the UK (3,4). Most people with CUD do not seek out professional treatment (5,6), with non-treatment seekers citing preference for informal treatment options and desire to be self-reliant (7,8). Despite this, there is little quantitative guidance on how an individual who uses cannabis can reduce their likelihood of developing CUD.

Previous research has identified factors that might increase an individual’s risk of developing symptoms of CUD. One increasingly recognised factor appears to be how cannabis is used, including aspects of use behaviour such as frequency and quantity, as well as type, and relatedly the potency of cannabis product used. Frequency of use has been found to have a dose-response relationship with risk for CUD, whereby increased frequency (for example, using daily vs weekly vs monthly) is associated with increased risk of CUD (9).

However, research indicates that other factors relating to use are involved in conferring risk for CUD, which are not always well quantified in research. For example, cannabis potency (% THC) has been increasing for several decades (10) and use of high potency cannabis is associated with an increased risk of negative outcomes, including CUD and adverse mental health (11,12). Furthermore, quantity of cannabis used has also been linked to increased risk of CUD (13–15), and like cannabis potency, provides important data that is not captured in studies investigating frequency of use alone.

Recent consensus from different expert collaborations has highlighted the importance of increased sensitivity of data collection around cannabis use (16). Collecting high-quality data around cannabis use that can be harmonised across research studies can help to accurately assess outcomes related to cannabis use, including CUD. Furthermore, increasing accuracy of measurement may result in more practical harm reduction advice for cannabis users hoping to reduce their risk of CUD or other harms. For example, researchers developing lower-risk cannabis use guidelines (17), reviewed previous evidence and concluded that a reduction in frequency of days per week, using lower-potency products as well as delaying onset of cannabis use until after the duration of puberty may reduce cannabis-related harms. However, there is currently a lack of evidence to propose specific guidance around quantity of cannabis use to reduce cannabis-related harms. Dose-related information (based on standard alcohol units, or standard drinks) is the cornerstone of alcohol harm reduction guidance in many countries around the world. For example in the UK people who drink alcohol are advised not to regularly consume more than 14 units of alcohol per week to reduce the risk of harm.

There are currently no such quantitative guidelines for safer levels of cannabis use. This presents a key unmet public health need, given increases in cannabis use, cannabis potency, and treatment need internationally (18).

Previous proposals to improve data collection of cannabis quantity and estimate risk thresholds include the ‘standard joint unit’ of 7mg of THC, a quantity chosen based on median quantities present in joints donated to researchers by people who use cannabis (19). The standard joint unit represents an improvement from relying on frequency-only measures, by directly considering quantity of THC consumed. Researchers have previously proposed 1 standard joint unit per day as a threshold for moderate risk of having cannabis-related problems (using the cannabis abuse screening test, CAST) in individuals with heavy cannabis use (14). However, the dose of THC used in a joint may vary between and within individuals (e.g. morning versus evening use). Additionally, many people use several methods of administration as well as different types of cannabis that vary in THC quantity, that would not be captured by only measuring the number of joints used. However, research using the standard joint unit provides proof-of-concept evidence for the use of a quantity-based measure (number of joints consumed) to discriminate risk of problematic cannabis use.

Other recent proposals include measuring cannabis use by the quantity of THC consumed across all cannabis products and administration methods. This is particularly relevant given the increasing diversity of different cannabis types including high potency concentrates that have been observed in newly legal markets (20,21). Research has indicated that increasing quantity of THC consumed is associated with an increased number of CUD symptoms (22). A standard “unit” of 5mg THC has been proposed (23), and implemented by the US National Insitutes of Health, who now mandate the use/reporting of THC units in research they fund (24). Using baseline data from the CannTeen study (a 12-month observational longitudinal dataset of adult and adolescent cannabis users) we recently validated a method of estimating standard THC units in observational studies, using an Enhanced Cannabis Timeline Followback (EC-TLFB; 22) Standard THC units provided the strongest correlation with urinary markers of cannabis use (creatinine-adjusted THC-COOH) of all cannabis use measures included. Following this validation, we now present evidence on the ability of standard THC units (estimated using the EC-TLFB, which was administered every three months for one year in the longitudinal CannTeen study) to discriminate risk of DSM-5 CUD. This paper represents what we believe is the first attempt to determine thresholds for risk of CUD by weekly standard THC units consumed.

The current study aimed to establish preliminary risk thresholds for CUD from standard THC units using receiver operating characteristic (ROC) curve analysis. We aimed to identify thresholds for *any* CUD (mild, moderate or severe), as well as greater severity, *at least moderate* CUD. The CannTeen study includes matched groups of adolescents (aged 16-17) and adults (aged 26-29). This is important because prevalence of cannabis use is particularly high in adolescent groups, who also are more vulnerable to development of CUD than adults or older age groups (1,26–28). Whilst the only way of ensuring no harm from cannabis is to not use at all (17), people who use cannabis could benefit from accurate information regarding their risk, particularly in high-risk groups (e.g., adolescents). Therefore, in the current study we used data from both adolescent (16-17) and adults (26-29) to estimate the ability of standard THC units to discriminate risk of CUD separately by age group. We hypothesised that all ROC curves would have an area under the curve (AUC) greater than 0.70, indicating a good discrimination (and significantly better than chance) of THC units on the classification of individuals as CUD/ no CUD.

## METHOD

### Cannabis research context

This study used data from CannTeen, a longitudinal study of adult and adolescents who use cannabis in London, UK, that ran from November 2017 to June 2021 (sessions after 23rd March 2020 adapted to virtual data collection during the national COVID-19 lockdown periods in the UK). To put our research into a wider context (29), recreational cannabis use in the UK is illegal, and possession and supply can result in legal penalties. Medicinal cannabis was legalised in 2018 however access remains very limited, and it can only be prescribed by specialist doctors in limited circumstances. Cannabis is the most used illicit drug in the UK, with an estimated 6.8% of the adult population, and 13.8% of the young adult population reporting use in the past year (30). Most people who use cannabis in the UK do so by mixing herbal cannabis (estimated at 14.2% potency) (29) with tobacco into “joints”(32), although administration methods and products do vary, as was evidenced in the current study.

### Study Population

Participants were recruited from the Greater London area through social media and Gumtree advertisements, school assemblies, posters, flyers, and word of mouth. Participants were originally recruited as either cannabis users (1-7 days per week, past-3-months), or controls (0-10 lifetime uses of cannabis, no use in past 3 months, no intention to use in next month). The longitudinal study consisted of five visits three months apart, over the course of one year. Past-year diagnosis of DSM-5 CUD was assessed at the final session. As the time course of this assessment (past year) was completely matched with the time course of drug use assessments at visits 2-5, any participant who used cannabis at least once over these sessions (2-5) was included (including people who were recruited into either the cannabis or control groups at baseline). The two age groups were adolescents (16-17 years old) and adults (26-29 years old), recorded at the screening/baseline visit. Additional inclusion criteria assessed at the screening/baseline can be found in the Supplementary Materials.

### Ethical Approval

Ethical approval was obtained for the CannTeen study from the University College London ethics committee, project ID 5929/003. This analysis received a favourable ethical approval from the University of Bath ethics committee, reference number 10499-1232. The study was performed in accordance with the ethical standards laid out in the 1964 Declaration of Helsinki and its later amendments. All participants gave written, informed consent prior to their inclusion in the study.

### Assessments

#### Outcome

The outcome in this analysis was a DSM-5 diagnoses of CUD, as assessed by the Mini International Neuropsychiatric Interview. This measure assesses the presence of 11 symptoms of CUD across the preceding 12-months. The number of symptoms met indicated disorder severity, 0-1 symptoms classified as *no* disorder, 2-3 symptoms classified as *mild*, 4-5 symptoms classified as *moderate*, and 6-11 symptoms classified as *severe*.

#### Predictor

The predictor in this analysis was weekly standard THC units, averaged across the 12-month period covering the past-year assessment for CUD symptoms. At each session participants provided detailed data on their cannabis use, including all types and methods of cannabis use over the preceding 3-month period, using the EC-TLFB procedure (25). For each method of cannabis use reported by a participant, the type of cannabis they used (sinsemilla, hash/resin, seeded herbal, other), their estimated quantity of cannabis product added to the method, and the amount personally consumed were recorded. Potency of the three main cannabis types listed as well as for ‘kief’ and ‘shatter’ was estimated based on the latest available data from the UK (31), see Table 1 for example conversions of different cannabis types and quantities to standard THC units. Validation of this method was provided in a previous publication (25). The number of occasions of use of each method was then recorded across the 3-month period. For each participant, their mean weekly THC units consumed was calculated across the period of their available data (see analysis plan for criteria related to missing data). This standard THC unit calculation approach was used in a previous study (33), but here we include all participants with any recorded cannabis use during the longitudinal study period, including those initially entering the studies as “controls”. Further details of how standard THC units were calculated, including in the presence of missing data, are provided in the supplementary materials.

**Table 1.** Example conversions to standard THC units from a median joint size (0.45g), based on median potencies reported in Potter et al. (2018)

|  | Potency (% THC) | Grams | Number of standard THC units |
| --- | --- | --- | --- |
| Strong herbal cannabis | 14.2 | 0.45 | 12.78 |
| Hash / resin | 6.3 | 0.45 | 5.67 |
| Weak seeded herbal cannabis | 3.5 | 0.45 | 3.78 |

### Analysis plan

All analyses were pre-registered on the Open Science Framework in April 2023 (https://osf.io/n796p). This analysis used ROC curve analysis, a widely used approach for investigating the ability of a test at distinguishing a binary classification. ROC curves plot the true positive and false positive rate of a test and from this, thresholds can be chosen to determine the best cut-off of the test in distinguishing individuals with and without CUD. AUC can also be calculated, which gives an indication of the general accuracy of the classification model. Classifications were no vs any CUD (including mild, moderate, and severe), and no vs moderate/severe CUD. The moderate/severe comparison was chosen to investigate indicators of higher severity of problems, considering the communication of different thresholds for levels of risk, whilst also maximising statistical power. Comparisons were run for adults and adolescents separately. Power analyses indicated that the sample size for all comparisons was sufficient to detect an AUC of 0.74, with 80% power compared to a null model of 0.50 (14), using the ratio of positive to negative cases in each comparison. To determine weekly THC unit thresholds, we selected thresholds that maximise sensitivity and specificity (i.e., Youden’s index). Outliers (5% and 95% percentile) were winsorized as specified in our pre-registration, due to the presence of implausibly high estimates of mean weekly THC units, as has been evidenced in our previous research as well as in other research studies (22,33,34). We also ran a sensitivity analysis using non-winsorized data to determine whether outliers impacted the model. This study is reported in accordance with the STROBE (Strengthening the Reporting of Observational Studies in Epidemiology) guidelines.

## RESULTS

Of 177 CannTeen participants who used cannabis over the 12 months, DSM-5 CUD diagnosis was completed for 162 participants at the final follow-up. Ten of these participants were excluded due to missing 2 or more follow-ups (not including the final follow-up). A further 2 participants were excluded due to lack of sufficient data to estimate their standard THC units (see Supplementary materials for more details). The final sample size included in each analysis were therefore as follows: no CUD (adult n = 30; adolescent n = 29), any CUD (adult n = 35; adolescent N = 56), moderate/severe CUD (adult n = 22; adolescent n = 44). See Table 2 for sample characteristics by age group.

**Table 1.**
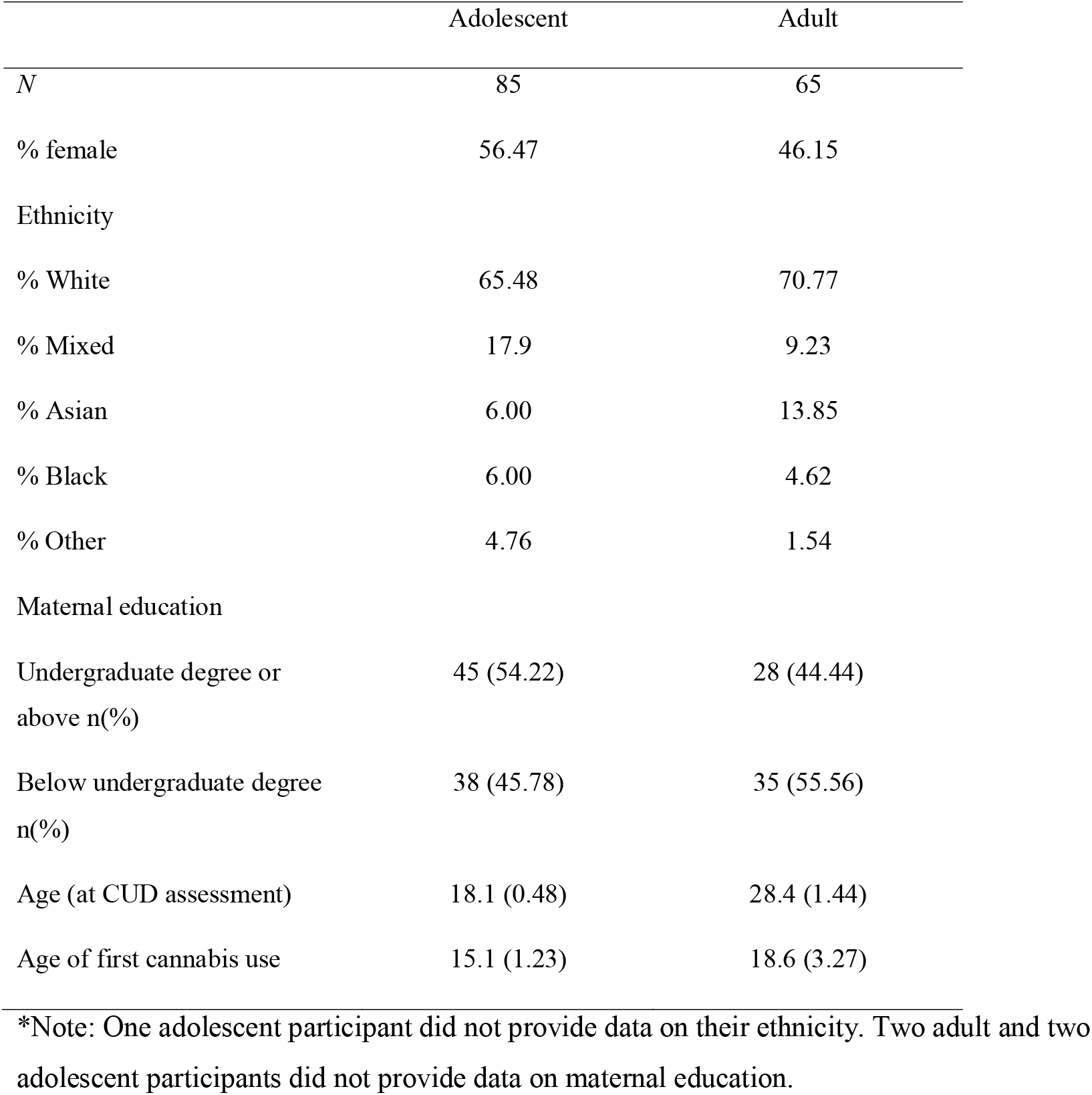
Sample characteristics. Data shown are frequencies and means (standard deviations) as appropriate.

|  | Adolescent | Adult |
| --- | --- | --- |
| *N* | 85 | 65 |
| % female | 56.47 | 46.15 |
| Ethnicity |  |  |
| % White | 65.48 | 70.77 |
| % Mixed | 17.9 | 9.23 |
| % Asian | 6.00 | 13.85 |
| % Black | 6.00 | 4.62 |
| % Other | 4.76 | 1.54 |
| Maternal education |  |  |
| Undergraduate degree or above n(%) | 45 (54.22) | 28 (44.44) |
| Below undergraduate degree n(%) | 38 (45.78) | 35 (55.56) |
| Age (at CUD assessment) | 18.1 (0.48) | 28.4 (1.44) |
| Age of first cannabis use | 15.1 (1.23) | 18.6 (3.27) |

\*Note: One adolescent participant did not provide data on their ethnicity. Two adult and two adolescent participants did not provide data on maternal education.

### No CUD vs any CUD

THC units provided a good discrimination of no CUD from *any* CUD (mild, moderate or severe), see *Figure 1a* and *1b*. The AUC for the adult model was acceptable, borderline excellent (0.79, 95%CIs: 0.68, 0.91), and for the adolescent model was outstanding (0.94, 95% CIs: 0.89, 0.99). For adults, the optimal threshold was 8.26 units per week (sensitivity = 0.86, specificity = 0.63), and for adolescents the optimal threshold was 6.04 units per week (sensitivity = 0.89, specificity = 0.90; see *Figure 2a* and *2b* and supplementary Tables 1 and 2 for full model outputs).

**Figure 1.**
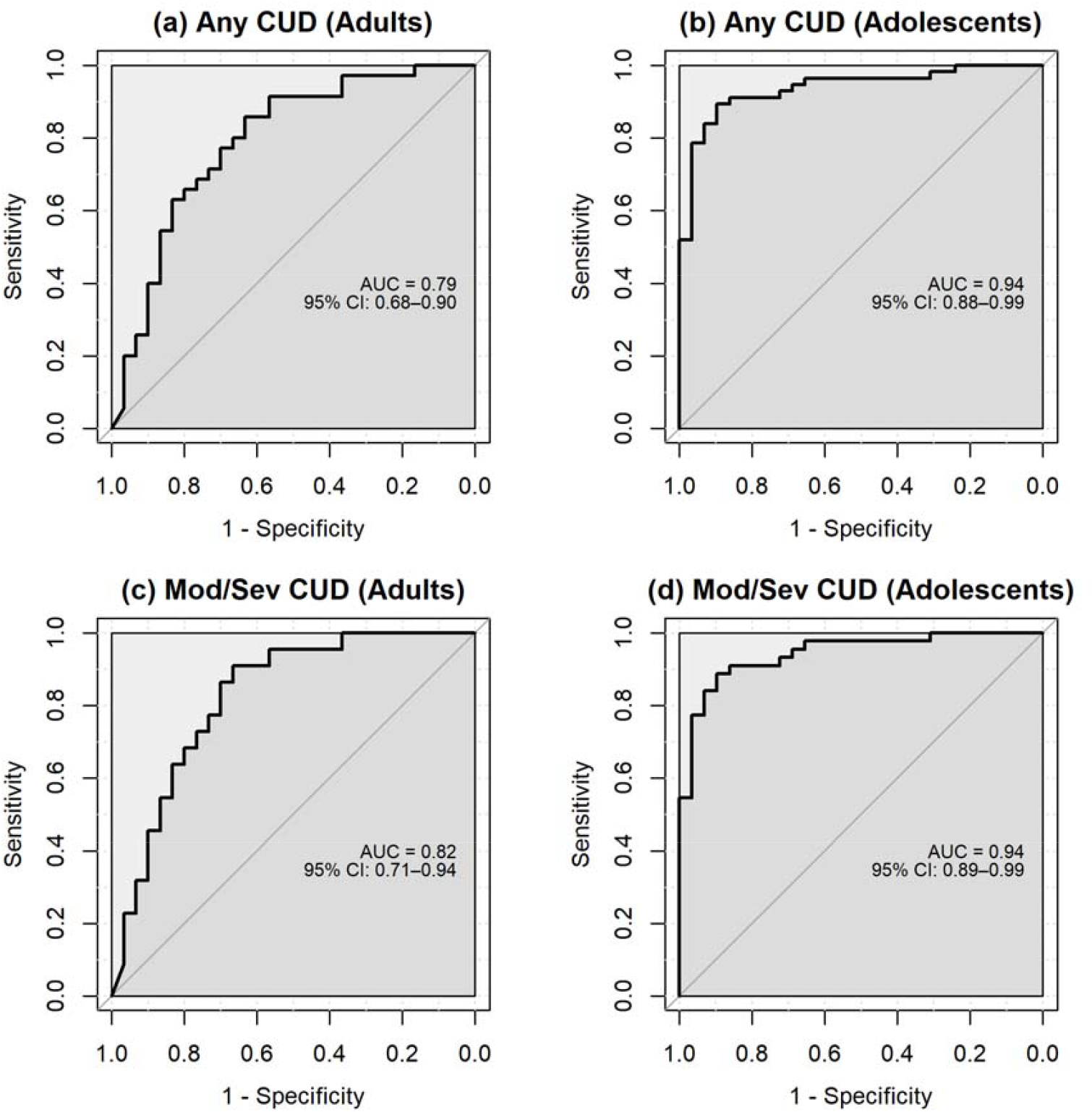
ROC curves for weekly standard THC units discriminating CUD and different severities

**Figure 2.**
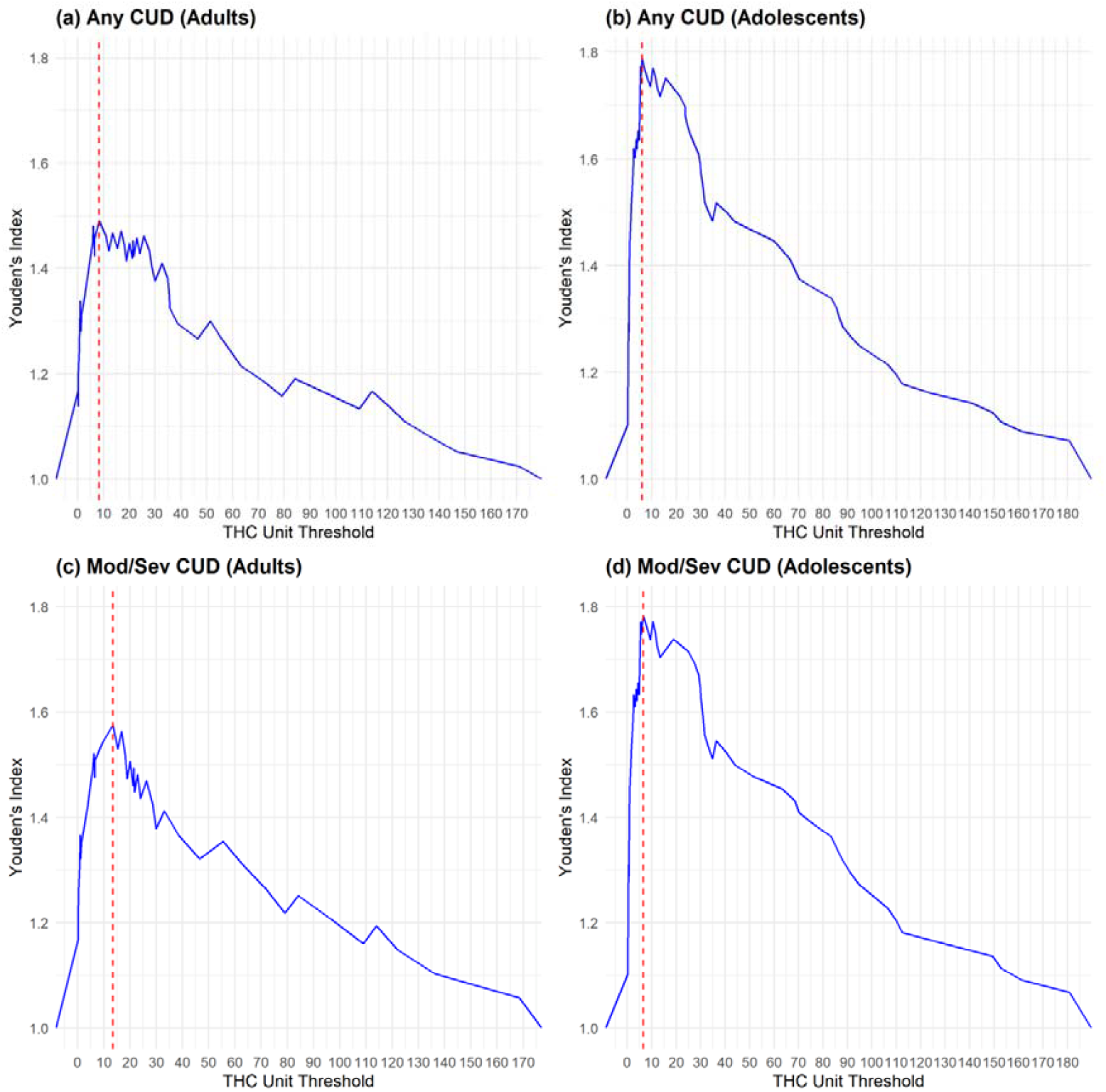
Youden’s Index (combined sensitivity and specificity) for weekly THC unit thresholds

### No CUD vs moderate/severe CUD

THC units provided a good discrimination of no CUD from *moderate/severe* CUD, see *Figure 1c* and *1d*. The AUC for the adult model was excellent (0.82, 95%CIs: 0.71, 0.94) and for the adolescent model was outstanding (0.94, 95%CIs: 0.89, 0.99). For adults, the optimal threshold was 13.44 units per week (sensitivity = 0.91, specificity = 0.67), whereas for adolescents the optimal threshold was 6.45 units per week (sensitivity = 0.89, specificity = 0.90; see *Figure 2c* and *2d*, and supplementary Tables 3 and 4).

### Sensitivity analysis

Sensitivity analyses confirmed that the pattern of results not materially different in the whole group sample vs the winsorized sample. The AUC of each model were consistent with the primary analyses, and the thresholds that maximised sensitivity and specificity were the same as those chosen in the main models using winsorized data. This indicates that potential outliers did not have a meaningful impact on the performance of the ROC models (see Supplementary Tables 5-8 and Supplementary Figures 1 and 2).

## DISCUSSION

Determining risk thresholds for CUD based on quantity of THC could inform harm-reduction strategies to reduce the health burden of CUD. To our knowledge, this is the first study to estimate risk thresholds for CUD based on standard THC units. In adults, 8.26 units per week was the optimal threshold for any CUD, and 13.44 units per week for moderate/severe CUD. In adolescents, the optimal thresholds were 6.04 units per week for any CUD, and 6.45 for moderate/severe CUD. Sensitivity of chosen thresholds across all models was high (86-91%), indicating that approximately 9 out of 10 individuals who used cannabis below the chosen weekly thresholds did not have CUD at the relevant severity. These preliminary findings may help to feed into the development of lower-risk guidelines for cannabis use, to aid cannabis users who wish to reduce their risk of harm by choosing to consume less THC than the above thresholds per week.

These findings are broadly comparable to the cut-off of one standard joint unit per day for moderate/severe CUD found previously (14), which translates to approximately 10 standard THC units per week. The current analysis builds on these prior findings by incorporating varied methods of cannabis administration (not joints alone), by measuring standard THC units via a comprehensive enhanced timeline follow back methodology and by using a clinical diagnosis of CUD (DSM-5), matched to the time course of past-year substance use assessment in this study.

One of the strengths of this study was its ability to estimate risk thresholds separately for adolescents (aged 16-17) and adults (aged 26-29). These analyses provided further support for a growing body of evidence showing that people who start use cannabis frequently during adolescence are at a greater risk of CUD compared to frequent use as adults (1,27,28,35). In the current analysis, the thresholds for lower risk were lower overall for adolescents than adults, indicating greater vulnerability to the effects of cannabis use on CUD risk.

Interestingly, when comparing the thresholds for different severities of CUD, among adolescents the thresholds for risk of any CUD and moderate/severe CUD were very similar, indicating that there was little to distinguish mild CUD from moderate/severe CUD in adolescents. In adults, a more typical dose-response relationship emerged, with approximately 1.5 times greater number of weekly THC units needed to confer risk for moderate/severe CUD than for any CUD. This could indicate that guidelines based on dose-related risk may be particularly useful for adults, given a strong dose-response relationship. For adolescents, the thresholds were low and a lack of dose-response relationship indicates that interventions to prevent/stop use completely, or keep use at very minimal levels (i.e. below 6.04 units per week) may be more appropriate. Reducing quantity of THC units consumed can be achieved by reducing the amount of cannabis used in each administration, using lower THC products, or reducing frequency (uses per day, days per week) of use.

As stated in current evidence-based lower risk cannabis use guidelines, the safest level of cannabis use is no use at all. This is particularly important when discussing the risks associated with different levels of cannabis use in populations who may be at heightened vulnerability to cannabis harms. In this study, we selected thresholds that maximised both total sensitivity and specificity. For most comparisons, there were other thresholds that showed similar levels of total sensitivity plus specificity, and generally there is a trade-off between sensitivity and specificity in these types of analyses. The model thresholds chosen here based on maximising sensitivity and specificity generally had lower specificity than sensitivity for adult models. For example, approximately 4 out of 10 people who used weekly standard THC units above the 8.26 threshold did not have CUD. Given that the application of these findings would be to communicate risk to prevent CUD, conservative thresholds that have high levels of sensitivity are important for providing guidance that would safely confer very little risk of CUD. In other settings, researchers might be interested in selecting thresholds that maximise specificity, for example in screening for those at the greatest treatment need. In the adolescent models, optimal thresholds had high sensitivity and specificity, with approximately 9 out of 10 individuals who used above the 6.04 threshold reporting CUD, and 9 out of 10 individuals who used below this threshold not reporting CUD.

Strengths of this study include the comprehensive EC-TLFB methodology, providing a direct measure of the quantity of drug used (e.g. THC) for all cannabis products and methods of administration, as well as the use of a DSM-5 clinical interview to assess CUD symptoms, which means that these thresholds are potentially clinically relevant. However, the study findings should be considered in the light of several limitations. Firstly, despite being powered to detect an effect of 0.76 or above across all comparisons, the sample size was modest and the findings warrant replication in a larger and more representative samples of people who use cannabis. Additionally, 12 participants who had used cannabis over the assessment period and completed the DSM-5 CUD assessment were not included in this analysis due to not attending more than one intermediary session (n=10) or providing insufficient data to estimate their standard THC units (n=2). We chose not to include these participants due to concern around extrapolation of 6-months of missing data potentially reducing the accuracy of measurement, something that is a key strength of this analysis.

Previous studies have evidenced missing data when estimating THC use (22), in line with the increased complexity of the necessary data collection.

We stratified analyses by age group given previous robust evidence of differing risks for CUD by current age. However, the presence of other risk factors for both younger onset cannabis use as well as CUD, for example mental health disorders, were not considered in the current analysis, and therefore individual risk for CUD may still differ according to personal profile of risk. Additionally, the data for this analysis comes from the CannTeen study, for which participants met specific inclusion criteria that means they may not represent the wider population of people who use cannabis. Specifically, those who were recruited as adult regular cannabis users reported no weekly use of cannabis (across a 3-month period or longer), before the age of 18. This may differentiate their risk from adults who use cannabis and initiated regular use at an earlier age. Additionally, CannTeen participants were not seeking or in treatment for CUD and therefore results may be different in clinical samples.

Finally, standard THC units require more estimation by the participant than simple reporting of frequency of use, and therefore are more time-consuming data to collect as well as potentially open to more error in estimation (e.g., grams of use). However, standard THC units showed the largest effect size association with objectively quantified THC consumption in urine of all cannabis use assessments (25). Repeating this analysis in data where quantity and potency of use might be better characterised or where CUD endorsement may differ (e.g. in jurisdictions where cannabis is legal) would be an interesting future avenue for this research. Furthermore, repetition in large, nationally representative datasets will be needed to establish risk thresholds that are appropriate for us in public health messaging. To conclude, using standard THC units we identified risk thresholds for *any* CUD and *moderate/severe* CUD, separately for adults and adolescents who used cannabis in the past year. These preliminary findings could feed into the development of low-risk cannabis use guidelines around quantity of cannabis use.

## Data Availability

The participants of this study did not give written consent for their data to be shared publicly, so the research supporting data are not available.

## Acknowledgements

We would like to thank all the CannTeen participants for giving up their time to participate in this study. We would also like to thank everyone who contributed to data collection as well as Sharinjeet Dhiman.

## Notes

**Primary funding** Primary funding for the CannTeen study was supported by a grant from the Medical Research Council (MRC; award number MR/P012728/1) to HVC and TPF. TPF is supported by a UKRI Future Leaders Fellowship (MR/Y017560/1). This work was supported in part by grant MR/N0137941/1 for the GW4 BIOMED MRC DTP, awarded to the Universities of Bath, Bristol, Cardiff, and Exeter from the Medical Research Council (MRC)/UKRI.

### Competing Interest Statement

The authors have declared no competing interest.

### Funding Statement

Primary funding for the CannTeen study was supported by a grant from the Medical Research Council (MRC; award number MR/P012728/1) to HVC and TPF. TPF is supported by a UKRI Future Leaders Fellowship (MR/Y017560/1). This work was supported in part by grant MR/N0137941/1 for the GW4 BIOMED MRC DTP, awarded to the Universities of Bath, Bristol, Cardiff, and Exeter from the Medical Research Council (MRC)/UKRI.

### Author Declarations

Ethical approval was obtained for the CannTeen study from the University College London ethics committee, project ID 5929/003. This analysis received a favourable ethical approval from the University of Bath ethics committee, reference number 10499-1232.

