## supplementary materials for "Estimating thresholds for risk of cannabis use disorder using standard THC units"

**Inclusion criteria**

Inclusion criteria for all groups included fluency in English, ability to come to central London for all testing sessions, normal or corrected-to-normal vision, and capacity to give informed consent. Exclusion criteria were history of diagnosed psychotic episode or disorder, illicit drug use (excluding nitrous oxide) >2 times per month, over the past-3-months, nitrous oxide use >1 day per week over the past-3-months, receiving of treatment for any mental health condition (including CUD) in the past month, currently daily use of a medication which is commonly psychotropic, (6) any mental or physical health condition deemed problematic by a medical doctor, and age-adjusted BMI <2nd or >99.6th percentile. The adult cannabis group were required to not have used cannabis on a weekly basis averaged over 3 months under the age of 18

**Calculating standard THC unit data**

Standard THC units were calculated according to the protocol of our previous work (1). Relevant data were collected using the Enhanced Cannabis Timeline Followback (2). At each assessment occasion, participants were asked questions about their cannabis use in the preceding 3-month period, listing all ‘types’ of cannabis used (including but not limited to strong herbal cannabis, hash/resin, weak seeded herbal, THC oil). For each product type, they indicated the methods that they had administrated cannabis with (including but not limited to joint, bong, pipe, edibles, vaporiser). For each cannabis type / administration combination, the participant indicated the typical amount of cannabis they would add to the method, and whether they would consume the whole amount themselves or how much they would share with others. Following the TLFB approach, participants would then report for each day of the preceding 3-month period whether they had used cannabis, and if so which type/administration combinations they had used. To convert this information into total grams of THC consumed, we used estimates of potency based on the latest available UK data (3), which were 14.2% for stronger herbal cannabis, 6.3% for hash/resin and 3.5% for weaker seeded herbal cannabis. The number of standard THC units in each method was calculated as the grams of cannabis personally consumed, divided by potency of product type, divided by 0.005 (the quantity of a standard THC unit, in grams). The number of THC units consumed was then summed to represent the total over the 3-month assessment period, which was then converted into a mean weekly score.

Participants were able to estimate the quantity of cannabis they had used in each method in the vast majority of cases. Occasionally, participants found it difficult or refused to estimate the grams of cannabis they used in a particular method. This occurred for methods such as edibles (including brownies, cookies, cake, chocolate, butter and sweets), where the participant may not have seen the amount of cannabis added to the method. In the first instance, if the participant had used this method at a different occasion and provided an estimate of quantity, we substituted this value for the missing value. In cases where the participant did not have other data to estimate from, we used average data from the whole CannTeen sample for each type. Median quantities of different edible types are found in Table 1.

Supplementary Table 1. Estimates of cannabis quantity across types of ‘edibles’

| Cannabis edible type | Median quantity (g) from whole sample |
| --- | --- |
| Sinsemilla cannabis in a brownie/cookie/cake | 0.45 |
| Sinsemilla cannabis in chocolate | 0.1 |
| Sinsemilla cannabis in butter | 0.5 |
| Sinsemilla cannabis in a sweet/gummy | 0.075 |

Participants also struggled to estimate quantity when reporting use of THC oil in a vaporiser. When data was provided for this method, it tended to be inconsistent, with some participants providing data on the quantity in a cartridge but estimating their uses in terms of ‘puffs’ or giving implausible estimates for one occasion of use (e.g., 20ml), that varied from other occasions. Because of this, we decided that using the whole group median (0.3g) was inappropriate, as doing so would very likely lead to overestimation, particularly in cases when use was reported as only one or two puffs. Instead, for all occasions that a participant reported THC use in a vaporiser, we estimated quantity by calculating the mean of the other cannabis administration methods used by the participant at that occasion, and substituted potency for the mean of the main three cannabis types (8%). Two CannTeen participants reported use of THC oil in a vaporiser as their primary method of use on several occasions, with inconsistent quantity estimates and inadequate data from other methods of cannabis use to estimate from. We decided that estimating quantity and potency of use for these participants was not feasible, and their data was not used in this analysis.

Finally, a small number of participants reported use of trichome powder ‘kief’ added on top of a sinsemilla joint but did not estimate the quantity, describing this as a ‘sprinkle’ and the amount as ‘negligible’. We consulted with a person who uses cannabis (expert by experience) who uses this method to get an estimate of the number of grams in a ‘sprinkle’ of trichome powder added to a joint. They estimated this to be 0.01 grams, which was substituted for the quantity in two cases.

.

### Thresholds for main model comparisons

Table 1. Any CUD – adults

| Threshold | Specificity | Sensitivity | Sensitivity + Specificity |
| --- | --- | --- | --- |
| -inf | 0 | 1 | 1 |
| 0.161 | 0.167 | 1.000 | 1.167 |
| 0.181 | 0.167 | 0.971 | 1.138 |
| 0.251 | 0.200 | 0.971 | 1.171 |
| 0.314 | 0.233 | 0.971 | 1.205 |
| 0.465 | 0.267 | 0.971 | 1.238 |
| 0.649 | 0.300 | 0.971 | 1.271 |
| 0.808 | 0.333 | 0.971 | 1.305 |
| 0.939 | 0.367 | 0.971 | 1.338 |
| 1.089 | 0.367 | 0.943 | 1.310 |
| 1.275 | 0.367 | 0.914 | 1.281 |
| 1.530 | 0.400 | 0.914 | 1.314 |
| 2.689 | 0.433 | 0.914 | 1.348 |
| 3.772 | 0.467 | 0.914 | 1.381 |
| 4.709 | 0.500 | 0.914 | 1.414 |
| 5.705 | 0.533 | 0.914 | 1.448 |
| 5.977 | 0.567 | 0.914 | 1.481 |
| 6.271 | 0.567 | 0.886 | 1.452 |
| 6.524 | 0.567 | 0.857 | 1.424 |
| 6.624 | 0.600 | 0.857 | 1.457 |
| *8.259 | 0.633 | 0.857 | 1.490 |
| 10.871 | 0.633 | 0.829 | 1.462 |
| 12.029 | 0.633 | 0.800 | 1.433 |
| 13.440 | 0.667 | 0.800 | 1.467 |
| 15.342 | 0.667 | 0.771 | 1.438 |
| 16.776 | 0.700 | 0.771 | 1.471 |
| 18.091 | 0.700 | 0.743 | 1.443 |
| 18.803 | 0.700 | 0.714 | 1.414 |
| 19.972 | 0.733 | 0.714 | 1.448 |
| 21.097 | 0.733 | 0.686 | 1.419 |
| 21.377 | 0.767 | 0.686 | 1.452 |
| 21.660 | 0.767 | 0.657 | 1.424 |
| 22.832 | 0.800 | 0.657 | 1.457 |
| 23.976 | 0.800 | 0.629 | 1.429 |
| 25.669 | 0.833 | 0.629 | 1.462 |
| 27.722 | 0.833 | 0.600 | 1.433 |
| 28.646 | 0.833 | 0.571 | 1.405 |
| 29.982 | 0.833 | 0.543 | 1.376 |
| 32.676 | 0.867 | 0.543 | 1.410 |
| 34.863 | 0.867 | 0.514 | 1.381 |
| 35.371 | 0.867 | 0.486 | 1.352 |
| 35.678 | 0.867 | 0.457 | 1.324 |
| 38.792 | 0.867 | 0.429 | 1.295 |
| 46.508 | 0.867 | 0.400 | 1.267 |
| 51.426 | 0.900 | 0.400 | 1.300 |
| 55.067 | 0.900 | 0.371 | 1.271 |
| 59.133 | 0.900 | 0.343 | 1.243 |
| 63.181 | 0.900 | 0.314 | 1.214 |
| 72.010 | 0.900 | 0.286 | 1.186 |
| 79.050 | 0.900 | 0.257 | 1.157 |
| 84.155 | 0.933 | 0.257 | 1.190 |
| 96.754 | 0.933 | 0.229 | 1.162 |
| 109.029 | 0.933 | 0.200 | 1.133 |
| 113.969 | 0.967 | 0.200 | 1.167 |
| 120.383 | 0.967 | 0.171 | 1.138 |
| 126.625 | 0.967 | 0.143 | 1.110 |
| 136.201 | 0.967 | 0.114 | 1.081 |
| 146.613 | 0.967 | 0.086 | 1.052 |
| 171.070 | 0.967 | 0.057 | 1.024 |
| Inf | 1 | 0 | 1 |

Table 2. Any CUD – adolescent

| Threshold | Specificity | Sensitivity | Sensitivity + Specificity |
| --- | --- | --- | --- |
| -inf | 0 | 1 | 1 |
| 0.180 | 0.103 | 1.000 | 1.103 |
| 0.210 | 0.138 | 1.000 | 1.138 |
| 0.298 | 0.172 | 1.000 | 1.172 |
| 0.386 | 0.207 | 1.000 | 1.207 |
| 0.418 | 0.241 | 1.000 | 1.241 |
| 0.443 | 0.241 | 0.982 | 1.224 |
| 0.466 | 0.276 | 0.982 | 1.258 |
| 0.538 | 0.310 | 0.982 | 1.292 |
| 0.609 | 0.310 | 0.964 | 1.275 |
| 0.636 | 0.345 | 0.964 | 1.309 |
| 0.722 | 0.379 | 0.964 | 1.344 |
| 0.853 | 0.414 | 0.964 | 1.378 |
| 0.923 | 0.448 | 0.964 | 1.413 |
| 1.007 | 0.483 | 0.964 | 1.447 |
| 1.291 | 0.517 | 0.964 | 1.482 |
| 1.671 | 0.552 | 0.964 | 1.516 |
| 2.042 | 0.586 | 0.964 | 1.550 |
| 2.379 | 0.621 | 0.964 | 1.585 |
| 2.599 | 0.655 | 0.964 | 1.619 |
| 3.105 | 0.655 | 0.946 | 1.602 |
| 3.635 | 0.690 | 0.946 | 1.636 |
| 3.837 | 0.690 | 0.929 | 1.618 |
| 4.295 | 0.724 | 0.929 | 1.653 |
| 4.822 | 0.724 | 0.911 | 1.635 |
| 5.011 | 0.759 | 0.911 | 1.669 |
| 5.092 | 0.793 | 0.911 | 1.704 |
| 5.194 | 0.828 | 0.911 | 1.738 |
| 5.344 | 0.862 | 0.911 | 1.773 |
| 5.595 | 0.862 | 0.893 | 1.755 |
| *6.039 | 0.897 | 0.893 | 1.789 |
| 6.733 | 0.897 | 0.875 | 1.772 |
| 7.983 | 0.897 | 0.857 | 1.754 |
| 9.397 | 0.897 | 0.839 | 1.736 |
| 10.511 | 0.931 | 0.839 | 1.770 |
| 11.433 | 0.931 | 0.821 | 1.752 |
| 12.191 | 0.931 | 0.804 | 1.735 |
| 13.272 | 0.931 | 0.786 | 1.717 |
| 15.626 | 0.966 | 0.786 | 1.751 |
| 18.638 | 0.966 | 0.768 | 1.733 |
| 21.756 | 0.966 | 0.750 | 1.716 |
| 23.536 | 0.966 | 0.732 | 1.698 |
| 23.727 | 0.966 | 0.714 | 1.680 |
| 24.531 | 0.966 | 0.696 | 1.662 |
| 25.770 | 0.966 | 0.679 | 1.644 |
| 27.533 | 0.966 | 0.661 | 1.626 |
| 29.198 | 0.966 | 0.643 | 1.608 |
| 29.761 | 0.966 | 0.625 | 1.591 |
| 30.093 | 0.966 | 0.607 | 1.573 |
| 30.642 | 0.966 | 0.589 | 1.555 |
| 31.078 | 0.966 | 0.571 | 1.537 |
| 31.547 | 0.966 | 0.554 | 1.519 |
| 33.129 | 0.966 | 0.536 | 1.501 |
| 34.868 | 0.966 | 0.518 | 1.483 |
| 36.356 | 1.000 | 0.518 | 1.518 |
| 40.617 | 1.000 | 0.500 | 1.500 |
| 43.875 | 1.000 | 0.482 | 1.482 |
| 51.753 | 1.000 | 0.464 | 1.464 |

Table 3. Moderate/severe CUD – adult

| Threshold | Specificity | Sensitivity | Sensitivity + Specificity |
| --- | --- | --- | --- |
| -inf | 0 | 1 | 1 |
| 0.178 | 0.167 | 1.000 | 1.167 |
| 0.251 | 0.200 | 1.000 | 1.200 |
| 0.314 | 0.233 | 1.000 | 1.233 |
| 0.465 | 0.267 | 1.000 | 1.267 |
| 0.649 | 0.300 | 1.000 | 1.300 |
| 0.808 | 0.333 | 1.000 | 1.333 |
| 0.939 | 0.367 | 1.000 | 1.367 |
| 1.142 | 0.367 | 0.955 | 1.321 |
| 1.530 | 0.400 | 0.955 | 1.355 |
| 2.689 | 0.433 | 0.955 | 1.388 |
| 3.772 | 0.467 | 0.955 | 1.421 |
| 4.709 | 0.500 | 0.955 | 1.455 |
| 5.705 | 0.533 | 0.955 | 1.488 |
| 6.187 | 0.567 | 0.955 | 1.521 |
| 6.524 | 0.567 | 0.909 | 1.476 |
| 6.624 | 0.600 | 0.909 | 1.509 |
| 9.417 | 0.633 | 0.909 | 1.542 |
| *13.440 | 0.667 | 0.909 | 1.576 |
| 15.342 | 0.667 | 0.864 | 1.530 |
| 16.776 | 0.700 | 0.864 | 1.564 |
| 18.091 | 0.700 | 0.818 | 1.518 |
| 18.803 | 0.700 | 0.773 | 1.473 |
| 19.972 | 0.733 | 0.773 | 1.506 |
| 21.097 | 0.733 | 0.727 | 1.461 |
| 21.377 | 0.767 | 0.727 | 1.494 |
| 21.660 | 0.767 | 0.682 | 1.448 |
| 22.832 | 0.800 | 0.682 | 1.482 |
| 23.976 | 0.800 | 0.636 | 1.436 |
| 26.175 | 0.833 | 0.636 | 1.470 |
| 28.646 | 0.833 | 0.591 | 1.424 |
| 29.982 | 0.833 | 0.545 | 1.379 |
| 33.088 | 0.867 | 0.545 | 1.412 |
| 38.485 | 0.867 | 0.500 | 1.367 |
| 46.508 | 0.867 | 0.455 | 1.321 |
| 55.492 | 0.900 | 0.455 | 1.355 |
| 63.181 | 0.900 | 0.409 | 1.309 |
| 72.010 | 0.900 | 0.364 | 1.264 |
| 79.050 | 0.900 | 0.318 | 1.218 |
| 84.155 | 0.933 | 0.318 | 1.252 |
| 96.754 | 0.933 | 0.273 | 1.206 |
| 109.029 | 0.933 | 0.227 | 1.161 |
| 113.969 | 0.967 | 0.227 | 1.194 |
| 122.102 | 0.967 | 0.182 | 1.148 |
| 136.201 | 0.967 | 0.136 | 1.103 |
| 168.514 | 0.967 | 0.091 | 1.058 |
| Inf | 1 | 0 | 1 |

Table 4. Moderate/severe CUD – adolescent

| Threshold | Specificity | Sensitivity | Sensitivity + Specificity |
| --- | --- | --- | --- |
| -inf | 0 | 1 | 1 |
| 0.180 | 0.103 | 1.000 | 1.103 |
| 0.210 | 0.138 | 1.000 | 1.138 |
| 0.298 | 0.172 | 1.000 | 1.172 |
| 0.386 | 0.207 | 1.000 | 1.207 |
| 0.418 | 0.241 | 1.000 | 1.241 |
| 0.466 | 0.276 | 1.000 | 1.276 |
| 0.538 | 0.310 | 1.000 | 1.310 |
| 0.609 | 0.310 | 0.977 | 1.288 |
| 0.636 | 0.345 | 0.977 | 1.322 |
| 0.722 | 0.379 | 0.977 | 1.357 |
| 0.853 | 0.414 | 0.977 | 1.391 |
| 0.923 | 0.448 | 0.977 | 1.426 |
| 1.007 | 0.483 | 0.977 | 1.460 |
| 1.291 | 0.517 | 0.977 | 1.495 |
| 1.671 | 0.552 | 0.977 | 1.529 |
| 2.042 | 0.586 | 0.977 | 1.563 |
| 2.379 | 0.621 | 0.977 | 1.598 |
| 2.599 | 0.655 | 0.977 | 1.632 |
| 3.105 | 0.655 | 0.955 | 1.610 |
| 3.635 | 0.690 | 0.955 | 1.644 |
| 3.837 | 0.690 | 0.932 | 1.621 |
| 4.295 | 0.724 | 0.932 | 1.656 |
| 4.822 | 0.724 | 0.909 | 1.633 |
| 5.011 | 0.759 | 0.909 | 1.668 |
| 5.092 | 0.793 | 0.909 | 1.702 |
| 5.194 | 0.828 | 0.909 | 1.737 |
| 5.344 | 0.862 | 0.909 | 1.771 |
| 5.595 | 0.862 | 0.886 | 1.748 |
| *6.453 | 0.897 | 0.886 | 1.783 |
| 7.983 | 0.897 | 0.864 | 1.760 |
| 9.397 | 0.897 | 0.841 | 1.737 |
| 10.511 | 0.931 | 0.841 | 1.772 |
| 11.433 | 0.931 | 0.818 | 1.749 |
| 12.191 | 0.931 | 0.795 | 1.726 |
| 13.272 | 0.931 | 0.773 | 1.704 |
| 18.744 | 0.966 | 0.773 | 1.738 |
| 24.942 | 0.966 | 0.750 | 1.716 |
| 27.533 | 0.966 | 0.727 | 1.693 |
| 29.198 | 0.966 | 0.705 | 1.670 |
| 29.761 | 0.966 | 0.682 | 1.647 |
| 30.093 | 0.966 | 0.659 | 1.625 |
| 30.642 | 0.966 | 0.636 | 1.602 |
| 31.078 | 0.966 | 0.614 | 1.579 |
| 31.547 | 0.966 | 0.591 | 1.556 |
| 33.129 | 0.966 | 0.568 | 1.534 |
| 34.868 | 0.966 | 0.545 | 1.511 |
| 36.356 | 1.000 | 0.545 | 1.545 |
| 40.617 | 1.000 | 0.523 | 1.523 |
| 43.875 | 1.000 | 0.500 | 1.500 |
| 51.753 | 1.000 | 0.477 | 1.477 |
| 63.092 | 1.000 | 0.455 | 1.455 |
| 68.364 | 1.000 | 0.432 | 1.432 |
| 70.303 | 1.000 | 0.409 | 1.409 |
| 76.336 | 1.000 | 0.386 | 1.386 |
| 83.324 | 1.000 | 0.364 | 1.364 |
| 85.574 | 1.000 | 0.341 | 1.341 |
| 88.146 | 1.000 | 0.318 | 1.318 |
| 91.135 | 1.000 | 0.295 | 1.295 |
| 94.768 | 1.000 | 0.273 | 1.273 |
| 100.703 | 1.000 | 0.250 | 1.250 |
| 106.554 | 1.000 | 0.227 | 1.227 |
| 109.821 | 1.000 | 0.205 | 1.205 |
| 112.484 | 1.000 | 0.182 | 1.182 |
| 130.591 | 1.000 | 0.159 | 1.159 |
| 149.399 | 1.000 | 0.136 | 1.136 |
| 152.793 | 1.000 | 0.114 | 1.114 |
| 161.280 | 1.000 | 0.091 | 1.091 |
| 180.712 | 1.000 | 0.068 | 1.068 |
| Inf | 1 | 0 | 1 |

### Thresholds for sensitivity comparisons

Table 5. Any CUD – adults (non-winsorized data)

| Threshold | Specificity | Sensitivity | Sensitivity + Specificity |
| --- | --- | --- | --- |
| -inf | 0 | 1 | 1 |
| 0.048 | 0.033 | 1.000 | 1.033 |
| 0.095 | 0.067 | 1.000 | 1.067 |
| 0.120 | 0.100 | 1.000 | 1.100 |
| 0.145 | 0.133 | 1.000 | 1.133 |
| 0.161 | 0.167 | 1.000 | 1.167 |
| 0.181 | 0.167 | 0.971 | 1.138 |
| 0.251 | 0.200 | 0.971 | 1.171 |
| 0.314 | 0.233 | 0.971 | 1.205 |
| 0.465 | 0.267 | 0.971 | 1.238 |
| 0.649 | 0.300 | 0.971 | 1.271 |
| 0.808 | 0.333 | 0.971 | 1.305 |
| 0.939 | 0.367 | 0.971 | 1.338 |
| 1.089 | 0.367 | 0.943 | 1.310 |
| 1.275 | 0.367 | 0.914 | 1.281 |
| 1.530 | 0.400 | 0.914 | 1.314 |
| 2.689 | 0.433 | 0.914 | 1.348 |
| 3.772 | 0.467 | 0.914 | 1.381 |
| 4.709 | 0.500 | 0.914 | 1.414 |
| 5.705 | 0.533 | 0.914 | 1.448 |
| 5.977 | 0.567 | 0.914 | 1.481 |
| 6.271 | 0.567 | 0.886 | 1.452 |
| 6.524 | 0.567 | 0.857 | 1.424 |
| 6.624 | 0.600 | 0.857 | 1.457 |
| *8.259 | 0.633 | 0.857 | 1.490 |
| 10.871 | 0.633 | 0.829 | 1.462 |
| 12.029 | 0.633 | 0.800 | 1.433 |
| 13.440 | 0.667 | 0.800 | 1.467 |
| 15.342 | 0.667 | 0.771 | 1.438 |
| 16.776 | 0.700 | 0.771 | 1.471 |
| 18.091 | 0.700 | 0.743 | 1.443 |
| 18.803 | 0.700 | 0.714 | 1.414 |
| 19.972 | 0.733 | 0.714 | 1.448 |
| 21.097 | 0.733 | 0.686 | 1.419 |
| 21.377 | 0.767 | 0.686 | 1.452 |
| 21.660 | 0.767 | 0.657 | 1.424 |
| 22.832 | 0.800 | 0.657 | 1.457 |
| 23.976 | 0.800 | 0.629 | 1.429 |
| 25.669 | 0.833 | 0.629 | 1.462 |
| 27.722 | 0.833 | 0.600 | 1.433 |
| 28.646 | 0.833 | 0.571 | 1.405 |
| 29.982 | 0.833 | 0.543 | 1.376 |
| 32.676 | 0.867 | 0.543 | 1.410 |
| 34.863 | 0.867 | 0.514 | 1.381 |
| 35.371 | 0.867 | 0.486 | 1.352 |
| 35.678 | 0.867 | 0.457 | 1.324 |
| 38.792 | 0.867 | 0.429 | 1.295 |
| 46.508 | 0.867 | 0.400 | 1.267 |
| 51.426 | 0.900 | 0.400 | 1.300 |
| 55.067 | 0.900 | 0.371 | 1.271 |
| 59.133 | 0.900 | 0.343 | 1.243 |
| 63.181 | 0.900 | 0.314 | 1.214 |
| 72.010 | 0.900 | 0.286 | 1.186 |
| 79.050 | 0.900 | 0.257 | 1.157 |
| 84.155 | 0.933 | 0.257 | 1.190 |
| 96.754 | 0.933 | 0.229 | 1.162 |
| 109.029 | 0.933 | 0.200 | 1.133 |
| 113.969 | 0.967 | 0.200 | 1.167 |
| 120.383 | 0.967 | 0.171 | 1.138 |
| 126.625 | 0.967 | 0.143 | 1.110 |
| 136.201 | 0.967 | 0.114 | 1.081 |
| 146.613 | 0.967 | 0.086 | 1.052 |
| 238.808 | 0.967 | 0.057 | 1.024 |
| 347.220 | 0.967 | 0.029 | 0.995 |
| 386.478 | 0.967 | 0.000 | 0.967 |
| Inf | 1 | 0 | 1 |

Table 6. Any CUD – adolescents (non-winsorized data)

| Threshold | Specificity | Sensitivity | Sensitivity + Specificity |
| --- | --- | --- | --- |
| -inf | 0 | 1 | 1 |
| 0.074 | 0.034 | 1.000 | 1.034 |
| 0.102 | 0.069 | 1.000 | 1.069 |
| 0.164 | 0.103 | 1.000 | 1.103 |
| 0.210 | 0.138 | 1.000 | 1.138 |
| 0.298 | 0.172 | 1.000 | 1.172 |
| 0.386 | 0.207 | 1.000 | 1.207 |
| 0.418 | 0.241 | 1.000 | 1.241 |
| 0.443 | 0.241 | 0.982 | 1.224 |
| 0.466 | 0.276 | 0.982 | 1.258 |
| 0.538 | 0.310 | 0.982 | 1.292 |
| 0.609 | 0.310 | 0.964 | 1.275 |
| 0.636 | 0.345 | 0.964 | 1.309 |
| 0.722 | 0.379 | 0.964 | 1.344 |
| 0.853 | 0.414 | 0.964 | 1.378 |
| 0.923 | 0.448 | 0.964 | 1.413 |
| 1.007 | 0.483 | 0.964 | 1.447 |
| 1.291 | 0.517 | 0.964 | 1.482 |
| 1.671 | 0.552 | 0.964 | 1.516 |
| 2.042 | 0.586 | 0.964 | 1.550 |
| 2.379 | 0.621 | 0.964 | 1.585 |
| 2.599 | 0.655 | 0.964 | 1.619 |
| 3.105 | 0.655 | 0.946 | 1.602 |
| 3.635 | 0.690 | 0.946 | 1.636 |
| 3.837 | 0.690 | 0.929 | 1.618 |
| 4.295 | 0.724 | 0.929 | 1.653 |
| 4.822 | 0.724 | 0.911 | 1.635 |
| 5.011 | 0.759 | 0.911 | 1.669 |
| 5.092 | 0.793 | 0.911 | 1.704 |
| 5.194 | 0.828 | 0.911 | 1.738 |
| 5.344 | 0.862 | 0.911 | 1.773 |
| 5.595 | 0.862 | 0.893 | 1.755 |
| *6.039 | 0.897 | 0.893 | 1.789 |
| 6.733 | 0.897 | 0.875 | 1.772 |
| 7.983 | 0.897 | 0.857 | 1.754 |
| 9.397 | 0.897 | 0.839 | 1.736 |
| 10.511 | 0.931 | 0.839 | 1.770 |
| 11.433 | 0.931 | 0.821 | 1.752 |
| 12.191 | 0.931 | 0.804 | 1.735 |
| 13.272 | 0.931 | 0.786 | 1.717 |
| 15.626 | 0.966 | 0.786 | 1.751 |
| 18.638 | 0.966 | 0.768 | 1.733 |
| 21.756 | 0.966 | 0.750 | 1.716 |
| 23.536 | 0.966 | 0.732 | 1.698 |
| 23.727 | 0.966 | 0.714 | 1.680 |
| 24.531 | 0.966 | 0.696 | 1.662 |
| 25.770 | 0.966 | 0.679 | 1.644 |
| 27.533 | 0.966 | 0.661 | 1.626 |
| 29.198 | 0.966 | 0.643 | 1.608 |
| 29.761 | 0.966 | 0.625 | 1.591 |
| 30.093 | 0.966 | 0.607 | 1.573 |
| 30.642 | 0.966 | 0.589 | 1.555 |
| 31.078 | 0.966 | 0.571 | 1.537 |
| 31.547 | 0.966 | 0.554 | 1.519 |
| 33.129 | 0.966 | 0.536 | 1.501 |
| 34.868 | 0.966 | 0.518 | 1.483 |
| 36.356 | 1.000 | 0.518 | 1.518 |
| 40.617 | 1.000 | 0.500 | 1.500 |
| 43.875 | 1.000 | 0.482 | 1.482 |
| 51.753 | 1.000 | 0.464 | 1.464 |
| 59.950 | 1.000 | 0.446 | 1.446 |
| 63.427 | 1.000 | 0.429 | 1.429 |
| 66.637 | 1.000 | 0.411 | 1.411 |
| 68.432 | 1.000 | 0.393 | 1.393 |
| 70.303 | 1.000 | 0.375 | 1.375 |
| 76.336 | 1.000 | 0.357 | 1.357 |
| 83.324 | 1.000 | 0.339 | 1.339 |
| 85.455 | 1.000 | 0.321 | 1.321 |
| 86.606 | 1.000 | 0.304 | 1.304 |
| 88.146 | 1.000 | 0.286 | 1.286 |
| 91.135 | 1.000 | 0.268 | 1.268 |
| 94.768 | 1.000 | 0.250 | 1.250 |
| 100.703 | 1.000 | 0.232 | 1.232 |
| 106.554 | 1.000 | 0.214 | 1.214 |
| 109.821 | 1.000 | 0.196 | 1.196 |
| 112.484 | 1.000 | 0.179 | 1.179 |
| 123.930 | 1.000 | 0.161 | 1.161 |
| 140.659 | 1.000 | 0.143 | 1.143 |
| 149.399 | 1.000 | 0.125 | 1.125 |
| 152.793 | 1.000 | 0.107 | 1.107 |
| 161.280 | 1.000 | 0.089 | 1.089 |
| 191.645 | 1.000 | 0.071 | 1.071 |
| 234.172 | 1.000 | 0.054 | 1.054 |
| 396.515 | 1.000 | 0.036 | 1.036 |
| 676.304 | 1.000 | 0.018 | 1.018 |
| Inf | 1 | 0 | 1 |

Table 7. Moderate/severe CUD – adults (non-winsorized data)

| Threshold | Specificity | Sensitivity | Sensitivity + Specificity |
| --- | --- | --- | --- |
| -inf | 0 | 1 | 1 |
| 0.048 | 0.033 | 1.000 | 1.033 |
| 0.095 | 0.067 | 1.000 | 1.067 |
| 0.120 | 0.100 | 1.000 | 1.100 |
| 0.145 | 0.133 | 1.000 | 1.133 |
| 0.178 | 0.167 | 1.000 | 1.167 |
| 0.251 | 0.200 | 1.000 | 1.200 |
| 0.314 | 0.233 | 1.000 | 1.233 |
| 0.465 | 0.267 | 1.000 | 1.267 |
| 0.649 | 0.300 | 1.000 | 1.300 |
| 0.808 | 0.333 | 1.000 | 1.333 |
| 0.939 | 0.367 | 1.000 | 1.367 |
| 1.142 | 0.367 | 0.955 | 1.321 |
| 1.530 | 0.400 | 0.955 | 1.355 |
| 2.689 | 0.433 | 0.955 | 1.388 |
| 3.772 | 0.467 | 0.955 | 1.421 |
| 4.709 | 0.500 | 0.955 | 1.455 |
| 5.705 | 0.533 | 0.955 | 1.488 |
| 6.187 | 0.567 | 0.955 | 1.521 |
| 6.524 | 0.567 | 0.909 | 1.476 |
| 6.624 | 0.600 | 0.909 | 1.509 |
| 9.417 | 0.633 | 0.909 | 1.542 |
| *13.440 | 0.667 | 0.909 | 1.576 |
| 15.342 | 0.667 | 0.864 | 1.530 |
| 16.776 | 0.700 | 0.864 | 1.564 |
| 18.091 | 0.700 | 0.818 | 1.518 |
| 18.803 | 0.700 | 0.773 | 1.473 |
| 19.972 | 0.733 | 0.773 | 1.506 |
| 21.097 | 0.733 | 0.727 | 1.461 |
| 21.377 | 0.767 | 0.727 | 1.494 |
| 21.660 | 0.767 | 0.682 | 1.448 |
| 22.832 | 0.800 | 0.682 | 1.482 |
| 23.976 | 0.800 | 0.636 | 1.436 |
| 26.175 | 0.833 | 0.636 | 1.470 |
| 28.646 | 0.833 | 0.591 | 1.424 |
| 29.982 | 0.833 | 0.545 | 1.379 |
| 33.088 | 0.867 | 0.545 | 1.412 |
| 38.485 | 0.867 | 0.500 | 1.367 |
| 46.508 | 0.867 | 0.455 | 1.321 |
| 55.492 | 0.900 | 0.455 | 1.355 |
| 63.181 | 0.900 | 0.409 | 1.309 |
| 72.010 | 0.900 | 0.364 | 1.264 |
| 79.050 | 0.900 | 0.318 | 1.218 |
| 84.155 | 0.933 | 0.318 | 1.252 |
| 96.754 | 0.933 | 0.273 | 1.206 |
| 109.029 | 0.933 | 0.227 | 1.161 |
| 113.969 | 0.967 | 0.227 | 1.194 |
| 122.102 | 0.967 | 0.182 | 1.148 |
| 136.201 | 0.967 | 0.136 | 1.103 |
| 236.252 | 0.967 | 0.091 | 1.058 |
| 347.220 | 0.967 | 0.045 | 1.012 |
| 386.478 | 0.967 | 0.000 | 0.967 |
| Inf | 1 | 0 | 1 |

Table 8. Moderate/severe CUD – adolescents (non-winsorized data)

| Threshold | Specificity | Sensitivity | Sensitivity + Specificity |
| --- | --- | --- | --- |
| -inf | 0 | 1 | 1 |
| 0.074 | 0.034 | 1.000 | 1.034 |
| 0.102 | 0.069 | 1.000 | 1.069 |
| 0.164 | 0.103 | 1.000 | 1.103 |
| 0.210 | 0.138 | 1.000 | 1.138 |
| 0.298 | 0.172 | 1.000 | 1.172 |
| 0.386 | 0.207 | 1.000 | 1.207 |
| 0.418 | 0.241 | 1.000 | 1.241 |
| 0.466 | 0.276 | 1.000 | 1.276 |
| 0.538 | 0.310 | 1.000 | 1.310 |
| 0.609 | 0.310 | 0.977 | 1.288 |
| 0.636 | 0.345 | 0.977 | 1.322 |
| 0.722 | 0.379 | 0.977 | 1.357 |
| 0.853 | 0.414 | 0.977 | 1.391 |
| 0.923 | 0.448 | 0.977 | 1.426 |
| 1.007 | 0.483 | 0.977 | 1.460 |
| 1.291 | 0.517 | 0.977 | 1.495 |
| 1.671 | 0.552 | 0.977 | 1.529 |
| 2.042 | 0.586 | 0.977 | 1.563 |
| 2.379 | 0.621 | 0.977 | 1.598 |
| 2.599 | 0.655 | 0.977 | 1.632 |
| 3.105 | 0.655 | 0.955 | 1.610 |
| 3.635 | 0.690 | 0.955 | 1.644 |
| 3.837 | 0.690 | 0.932 | 1.621 |
| 4.295 | 0.724 | 0.932 | 1.656 |
| 4.822 | 0.724 | 0.909 | 1.633 |
| 5.011 | 0.759 | 0.909 | 1.668 |
| 5.092 | 0.793 | 0.909 | 1.702 |
| 5.194 | 0.828 | 0.909 | 1.737 |
| 5.344 | 0.862 | 0.909 | 1.771 |
| 5.595 | 0.862 | 0.886 | 1.748 |
| *6.453 | 0.897 | 0.886 | 1.783 |
| 7.983 | 0.897 | 0.864 | 1.760 |
| 9.397 | 0.897 | 0.841 | 1.737 |
| 10.511 | 0.931 | 0.841 | 1.772 |
| 11.433 | 0.931 | 0.818 | 1.749 |
| 12.191 | 0.931 | 0.795 | 1.726 |
| 13.272 | 0.931 | 0.773 | 1.704 |
| 18.744 | 0.966 | 0.773 | 1.738 |
| 24.942 | 0.966 | 0.750 | 1.716 |
| 27.533 | 0.966 | 0.727 | 1.693 |
| 29.198 | 0.966 | 0.705 | 1.670 |
| 29.761 | 0.966 | 0.682 | 1.647 |
| 30.093 | 0.966 | 0.659 | 1.625 |
| 30.642 | 0.966 | 0.636 | 1.602 |
| 31.078 | 0.966 | 0.614 | 1.579 |
| 31.547 | 0.966 | 0.591 | 1.556 |
| 33.129 | 0.966 | 0.568 | 1.534 |
| 34.868 | 0.966 | 0.545 | 1.511 |
| 36.356 | 1.000 | 0.545 | 1.545 |
| 40.617 | 1.000 | 0.523 | 1.523 |
| 43.875 | 1.000 | 0.500 | 1.500 |
| 51.753 | 1.000 | 0.477 | 1.477 |
| 63.092 | 1.000 | 0.455 | 1.455 |
| 68.364 | 1.000 | 0.432 | 1.432 |
| 70.303 | 1.000 | 0.409 | 1.409 |
| 76.336 | 1.000 | 0.386 | 1.386 |
| 83.324 | 1.000 | 0.364 | 1.364 |
| 85.574 | 1.000 | 0.341 | 1.341 |
| 88.146 | 1.000 | 0.318 | 1.318 |
| 91.135 | 1.000 | 0.295 | 1.295 |
| 94.768 | 1.000 | 0.273 | 1.273 |
| 100.703 | 1.000 | 0.250 | 1.250 |
| 106.554 | 1.000 | 0.227 | 1.227 |
| 109.821 | 1.000 | 0.205 | 1.205 |
| 112.484 | 1.000 | 0.182 | 1.182 |
| 130.591 | 1.000 | 0.159 | 1.159 |
| 149.399 | 1.000 | 0.136 | 1.136 |
| 152.793 | 1.000 | 0.114 | 1.114 |
| 161.280 | 1.000 | 0.091 | 1.091 |
| 191.645 | 1.000 | 0.068 | 1.068 |
| 234.172 | 1.000 | 0.045 | 1.045 |
| 533.296 | 1.000 | 0.023 | 1.023 |
| Inf | 1 | 0 | 1 |

### ROC curves for sensitivity comparisons

Supplementary Figure 1. None vs any CUD (non-winsorized data)

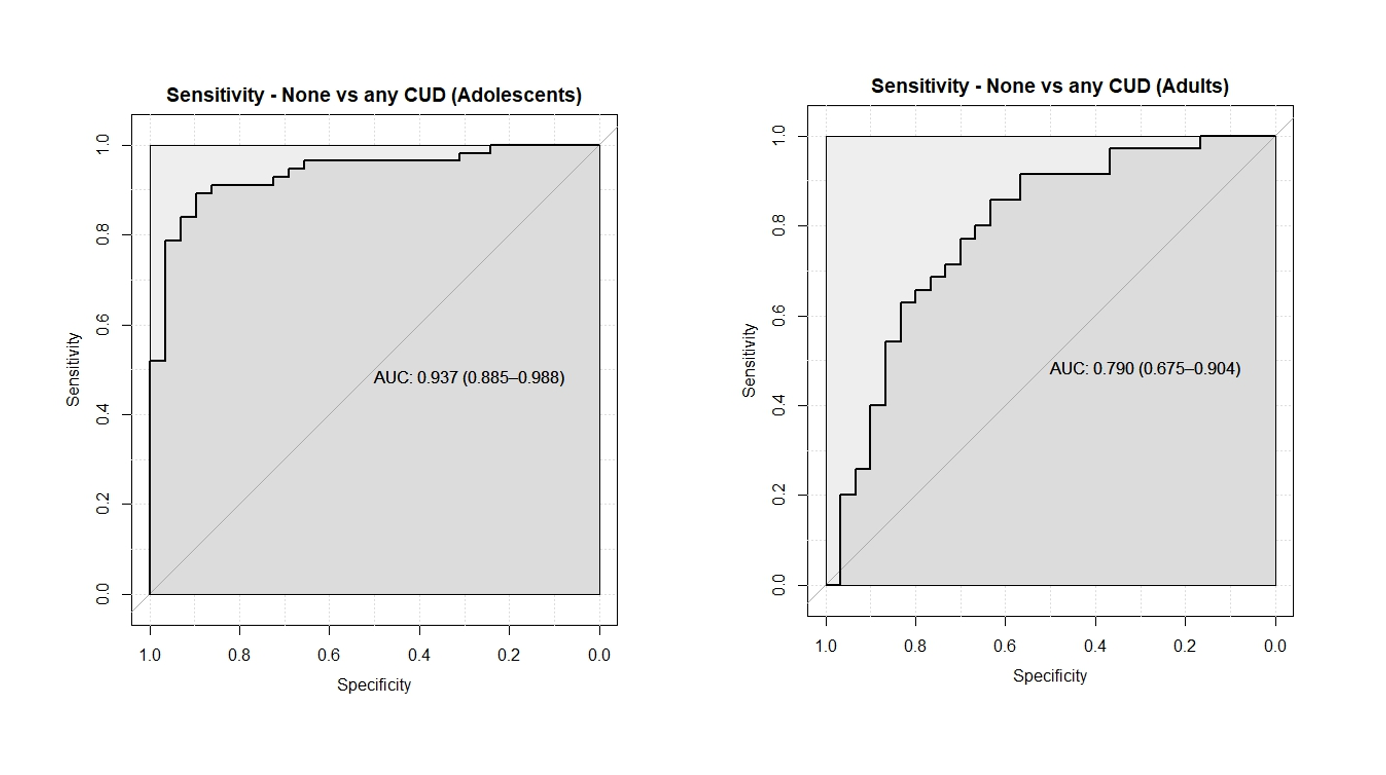

Supplementary Figure 2. None vs moderate/severe CUD (non-winsorized data)
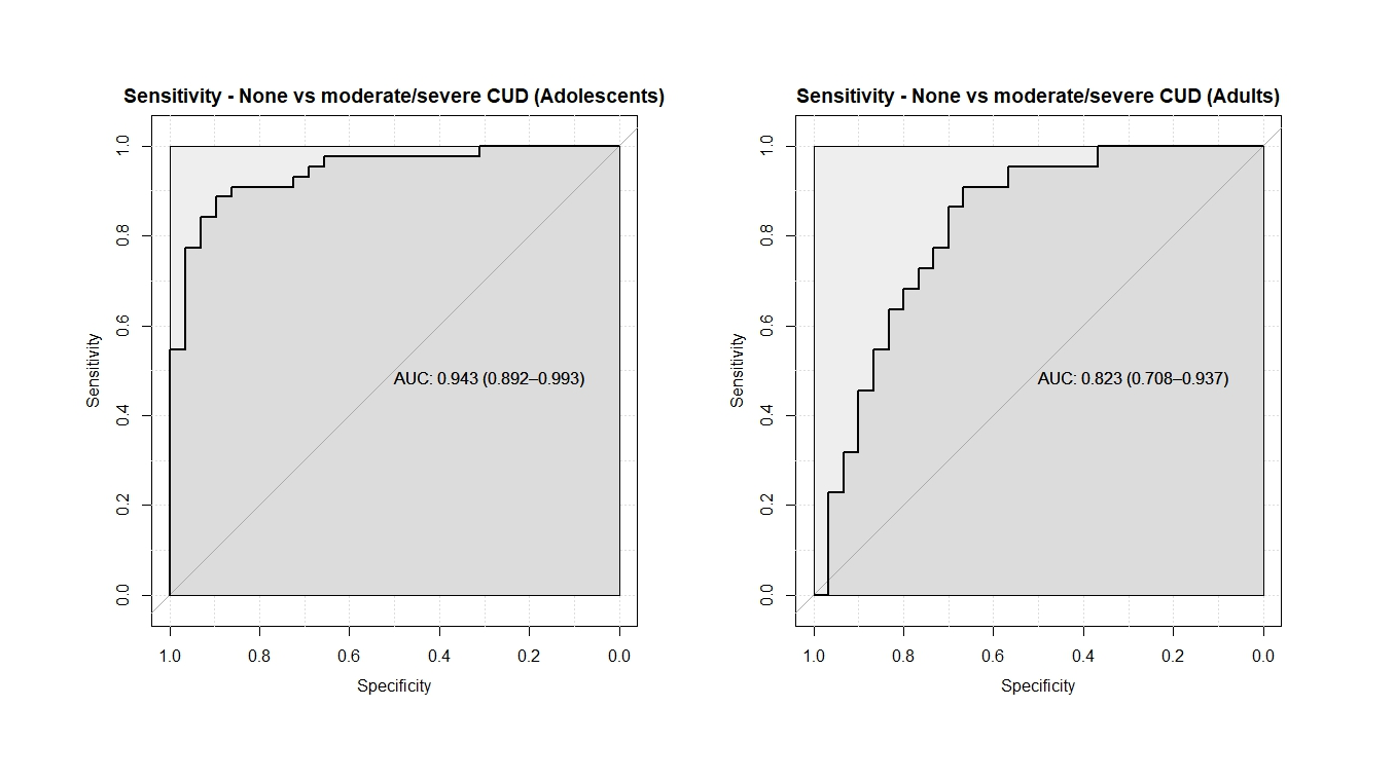
